# Development of a federated learning approach to predict acute kidney injury in adult hospitalized patients with COVID-19 in New York City

**DOI:** 10.1101/2021.07.25.21261105

**Authors:** Suraj K Jaladanki, Akhil Vaid, Ashwin S Sawant, Jie Xu, Kush Shah, Sergio Dellepiane, Ishan Paranjpe, Lili Chan, Patricia Kovatch, Alexander W Charney, Fei Wang, Benjamin S Glicksberg, Karandeep Singh, Girish N Nadkarni

## Abstract

Federated learning is a technique for training predictive models without sharing patient-level data, thus maintaining data security while allowing inter-institutional collaboration. We used federated learning to predict acute kidney injury within three and seven days of admission, using demographics, comorbidities, vital signs, and laboratory values, in 4029 adults hospitalized with COVID-19 at five sociodemographically diverse New York City hospitals, between March-October 2020. Prediction performance of federated models was generally higher than single-hospital models and was comparable to pooled-data models. In the first use-case in kidney disease, federated learning improved prediction of a common complication of COVID-19, while preserving data privacy.

## Introduction

Machine Learning (ML) models have been developed to aid in risk-stratification in patients with Coronavirus Disease-19 (COVID-19).^1-3^ Due to concerns about data privacy and ownership, most of these models were trained on data from single institutions, which limits their generalizability.^4, 5^ However, it is critical that these models be trained on large, diverse datasets, reflecting the variability seen in clinical practice.

Federated learning (FL), a technique that allows model training to occur without sharing confidential patient data, has gained significant interest during the pandemic. FL avoids the need to collect and store private data at a centralized location by allowing institutions to download a preliminary ML model, refine it locally, and then upload updated model parameters to an aggregator.^5-7^ FL models have been utilized to identify COVID-19 through computed tomography scans and to predict clinical outcomes in COVID-19 patients — published reports indicate that FL models not only outperform those trained on single-institution data but also approach the performance of traditional ML models trained on data pooled from multiple institutions.^6, 8-10^

COVID-19 has diverse clinical manifestations, and a common complication in hospitalized COVID-19 patients is acute kidney injury (AKI).^11-14^ Studies have reported AKI prevalence up to 46%, and mortality rates in AKI cohorts vary from 35%–71%.^15, 16^ Clinical models to pre-emptively identify patients with COVID-19 at increased risk for developing AKI can be valuable during a pandemic when resources may be constrained.^17^

Using COVID-19 associated AKI as a use case, we aimed to evaluate how FL trained models compare to models trained at individual institutions and pooled multi-institutional data. We developed federated models to predict AKI within three and seven days of admission using electronic health records (EHR) in patients hospitalized with COVID-19 and compared their performance to local and pooled models.

## Short Methods

The study population consisted of adult patients admitted to one of five Mount Sinai Health System hospitals in New York City with laboratory confirmed SARS-CoV-2 infection within 48 hours of admission between March 1^st^ and October 18^th^, 2020. We excluded patients with a history of transplants, a diagnosis of kidney failure or admissions less than 48 hours. We defined AKI according to 2012 Kidney Disease Improving Global Outcomes (KDIGO) guidelines.^18^ The primary outcomes of interest were incident AKI within three (AKI_3_) and seven (AKI_7_) days of admission. We included demographics, comorbidities, vital signs, laboratory values, and clinical outcomes extracted from an EHR database. For each parameter, we used the first available values obtained within 48 hours after admission. We used two classifiers: multilayer perceptron (MLP) to represent deep learning and Least Absolute Shrinkage and Selection Operator (LASSO) as a regression method. We used three training strategies for each classifier: local, federated, and pooled (Figure 1A, 1B). Local models for each hospital were trained and tested using only data from that hospital. Federated models used a FL framework to share model parameters with a central aggregator. Finally, pooled models combined data from all hospitals for model development and represented an optimal scenario. Model performance was assessed by average areas under the receiver operating characteristic curve (AUROCs) across 100 independent experiments with 70%/30% training/testing splits. SHapley Additive exPlanations (SHAP) scores were used to evaluate the importance of features contributing to model predictions.^19^ Detailed inclusion and exclusion criteria, data processing, classifier parameters, and software are described in Detailed Methods (Supplementary Material). We used the Transparent Reporting of a Multivariable Prediction Model for Individual Prognosis or Diagnosis (TRIPOD) guidelines and shared our code under the GNU General Public License v3 to enhance replicability (Supplementary Material).

**Figure 1.**
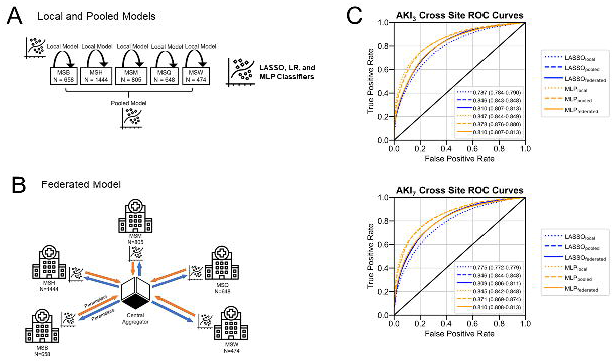
Study Design and Model Performance. (A) Overview of local and pooled models. Local models only utilize data from the site itself while pooled models incorporate data from all sites. Both local and pooled MLP, LR, and LASSO models were utilized. Based on https://medinform.jmir.org/2021/1/e24207/ (B) Overview of federated model. Parameters from a central aggregator are shared with each site, and sites do not have direct access to clinical data from others. After models are trained locally at a site, parameters are sent back to the central aggregator to update federated model parameters. Federated LASSO and MLP models were utilized. Based on https://medinform.jmir.org/2021/1/e24207/ (C) Performance of all models averaged across all sites as measured by area under the receiver-operating characteristic (AUROC) after 70-30 train-test split over 100 experiments with 95% confidence intervals for predicting AKI within three (top) and seven (bottom) days of admission.

## Results

### Cohort Characteristics

Among 4029 patients, the prevalence of AKI_3_ ranged from 23% to 37% across five hospitals, and 27% to 44% for AKI_7_. There were significant differences in demographics, comorbidities, laboratory measurements, outcome prevalence, and sample sizes between hospitals (Supplementary Table S3). For example, Mount Sinai West (MSW) had the largest class imbalance and smallest sample size of all hospitals.

### Learning Framework Comparisons

We evaluated performance of three model strategies (local, pooled, and federated) at each hospital for predicting AKI_3_ and AKI_7._ Performance averaged across the five sites is shown in Figure 1C. All models had AUROCs in the range of 0.70–0.90 for predicting AKI_3_ and AKI_7_. Pooled models consistently outperformed both local and federated models for AKI_3_ and AKI_7_ prediction. LASSO_federated_ also outperformed LASSO_local_ at all hospitals except one for both AKI_3_ and AKI_7_ prediction (Supplementary Table S5). MLP_federated_ outperformed MLP_local_ only at MSW for both AKI_3_ and AKI_7_ prediction (Supplementary Table S5). Additional performance metrics for all models are available in the supplementary material (Supplementary Table S4).

### MSW Performance and Feature Importance

The largest improvements in performance between local and federated models were seen for MSW (Figure 2A), with an increase of at least 3% (0.03) in AUROC for all three classifiers for AKI_3_ and AKI_7_ prediction. SHAP plots for LASSO_local_ and LASSO_federated_ models for AKI_3_ prediction at MSW demonstrate differences in feature importance (Figure 2B). For LASSO_local_, history of stroke, black race, and Hispanic/latino ethnicity were the three most important features for predicting AKI_3_. However, for the corresponding LASSO_federated_ model, the three features with highest importance were blood urea nitrogen, age, and albumin.

**Figure 2.**
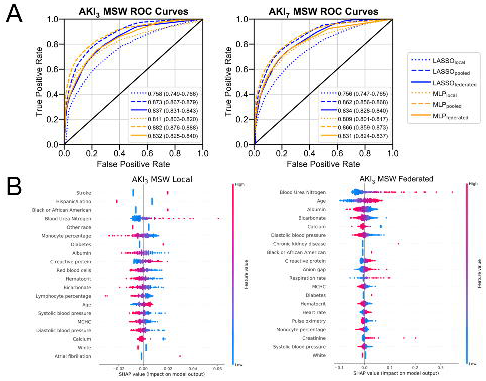
Model Performance and Feature Importance at MSW. (A) Performance of all models at Mount Sinai West (MSW) (n=474) as measured by area under the receiver-operating characteristic (AUROC) after 70-30 train-test split over 100 experiments with 95% confidence intervals for predicting AKI within three (left) and seven (right) days of admission. (B) SHAP values were calculated for LASSO_local_ (left) and LASSO_federated_ (right) for predicting AKI within three days of admission at MSW and illustrated in summary plots where features are listed in decreasing order of importance.

## Discussion

We trained models using federated learning to predict AKI at three and seven days in hospitalized patients with COVID-19. To our knowledge, this is the first use case of federated learning in kidney disease and to predict AKI over multiple time horizons. We found that models trained with FL outperformed those trained on locally available data, which was most apparent at the hospital with the smallest dataset.

All models had AUROCs of at least 0.70, suggesting that early vital signs and laboratory measurements can predict kidney complications during the first week in patients hospitalized with COVID-19. A similar approach has been used to predict mortality in hospitalized patients with COVID-19, highlighting the utility of peri-admission measurements in the prediction of clinical outcomes in this cohort.^10^ Model performance was consistently lower for AKI_7_ than AKI_3_, likely due to the longer prediction interval in the former case.

We found that LASSO_federated_ models outperformed their respective LASSO_local_ models at most hospitals. At MSW, the hospital with the smallest sample size and largest class imbalance, SHAP plots indicate that the LASSO_federated_ model attached greater importance to features that have more biologically plausible links to renal outcomes than the corresponding LASSO_local_ model, which was strongly influenced by race and a history of stroke. MSW was also the only site where MLP_federated_ outperformed MLP_local_, indicating that the greatest benefits from using FL to train deep learning models may accrue to sites with small amounts of local data available for training.

Models developed during this study may be implemented in clinical practice to predict onset of AKI in COVID-19 patients several days before it occurs, allowing clinicians to triage patients for better monitoring and use of intravenous fluids. Early prediction of AKI also allows for proactive allocation of resources, because published data indicate that these patients are hospitalized longer, are more likely to need intensive care, and more likely to require inpatient dialysis.^14^

Our study has several limitations. First, our data was limited to a single hospital system within New York City, which may limit generalizability to other geographical regions. Similarly, as the standard of care for hospitalized patients with COVID-19 has evolved after our data was collected, model performance should be validated with newer cohorts. Our models did not use inputs such as imaging and echocardiograms, and their exclusion may have hindered performance. We deliberately limited hyperparameter tuning of models to simulate a scenario where a federated model has to be urgently deployed at resource-constrained hospitals, such as during the COVID-19 pandemic. Allowing further optimization of the MLP structure, and techniques such as transfer learning may improve MLP_federated_ model performance.^20^ We examined only two classifiers; other approaches such as random forests or support vector machines may have yielded better results. Lastly, performance with the FL approach was generally statistically inferior to pooled dataset performance, and it remains to be explored if other approaches exist which can help bridge this gap while retaining the privacy advantages of FL.

In summary, we demonstrate the utility of FL to improve prediction of key outcomes while maintaining privacy and confidentiality. We hope this will encourage the development of generalizable clinical models which would otherwise be hindered by inability to share patient-level data across institutional boundaries.

## Data Availability

This article is written following the TRIPOD (Transparent Reporting of a Multivariable Prediction Model for Individual Prognosis or Diagnosis) guidelines, which are further elaborated in Supplementary Table 3. Furthermore, we release all code used for building the classifier under the GPLv3 license in a public GitHub repository.

https://github.com/HPIMS/COVID_Federated_AKI

