## Supplementary Text, Figures, Tables 1-2 for "Development of a federated learning approach to predict acute kidney injury in adult hospitalized patients with COVID-19 in New York City"

**Detailed Methods**

Study Population

The study population consisted of patients, aged 18 and over, admitted to one of five Mount Sinai Health System hospitals in New York City with laboratory confirmed SARS-CoV-2 infection within 48 hours of admission between March 1^st^ and October 18^th^, 2020. The five hospitals were Mount Sinai Brooklyn (MSB), Mount Sinai Hospital (MSH), Mount Sinai Morningside (MSM), Mount Sinai Queens (MSQ), and Mount Sinai West (MSW). This study was approved by the Institutional Review Board. We excluded patients with admissions of less than 48 hours’ duration, with a history of transplants, or diagnosis of end stage kidney disease (ESKD), identified by a combination of ESKD diagnosis and dialysis procedure International Classification of Diseases codes (Supplemental Figure S1). Patients presenting with AKI were included in the study. Further exclusion criteria included missing laboratory values and vital signs after admission.

Definition of Acute Kidney Injury

We defined AKI according to the 2012 Kidney Disease Improving Global Outcomes (KDIGO) guidelines, as an increase in serum creatinine of at least 0.3 mg/dL over 48 hours or 1.5 times baseline criteria.^1^ The most recent creatinine value during the time period between 365 and 7 days prior to admission was used as the baseline measurement. Missing baseline creatinine values were estimated using the Modification of Diet in Renal Disease Study equation assuming that baseline estimated GFR was 75 mL/min per 1.73 square meters of body surface area.^1^

Data Collection and Processing

Study data included demographics, comorbidities, vital signs, laboratory values, and clinical outcomes which were extracted from an EHR database. For each parameter, we used the first available values obtained within 48 hours after admission. We calculated medians and interquartile ranges for continuous data. Features with less than 70% missingness at each site were excluded, and we used k-Nearest Neighbors (k =5) to impute missing data for these features. Outliers below 0.5 or above 99.5 percentiles were excluded. We defined statistical significance as Bonferroni-adjusted *P* < 0.05 of Kruskal-Wallis or chi-squared tests.

We used the TRIPOD (Transparent Reporting of a Multivariable Prediction Model for Individual Prognosis or Diagnosis) guidelines to enhance replicability (Table S1). All code used to build our classifiers will be released under the GNU General Public License version 3 in a publicly accessible repository at <https://github.com/HPIMS/COVID_Federated_AKI>.

Study Design

We developed three model strategies for experiments: local, federated, and pooled. Local models were trained and tested on each hospital individually. Federated models used a federated learning (FL) framework to only share model parameters between hospitals. Finally, pooled models combined all hospital data for model training and testing and represented an optimal scenario where data privacy was not a concern. The primary outcomes of interest were AKI within three (AKI_3_) and seven (AKI_7_) days of admission.

Model Development

We developed two classifiers: multilayer perceptron (MLP) and Least Absolute Shrinkage and Selection Operator (LASSO). Each classifier had the same architecture for all hospitals to allow for fair comparisons. Limited hyperparameter tuning was performed to simulate a quick deployment scenario for ML models and each classifier had the same parameters for all hospitals to allow for fair comparisons. Final hyperparameters for classifiers are provided in Supplementary Table S2.

Our primary focus for this study was the performance of the FL models. To mimic a FL scenario, data was stored in isolated locations and model parameters were sent to a centralized location which represented the cloud. Model parameters include weights and biases learned during the training process. FL models were initialized with random parameters at the central aggregator and sent to each site for training for one epoch. After each epoch, model parameters were sent back to the aggregator, and federated averaging was performed. Federated averaging is a technique where model parameters are adjusted using the number of available data points and sums parameters for each layer.^2, 3^ Updated parameters after federated averaging were then sent to each site, and this process was repeated for a total of 80 epochs.

SHAP scores are generated through a game-theoretic approach and are often used for machine learning model interpretation. SHAP values were calculated for each feature and illustrated in summary plots where features are listed in decreasing order of importance.^4^

Differential Privacy and Gaussian Noise

Although federated learning is more secure than traditional pooled models as raw patient data do not leave their respective locations, parameters are still shared. To prevent inferring important properties of patient data from these shared parameters, differential privacy was incorporated into the MLP federated models to assess performance before and after the introduction of noise ^5-7^. We injected gaussian noise into the locally trained models before they were shared to the central location to achieve differential privacy (Supplemental Figures S2-S3).

**Statistical Analysis**

All models were built on Python 3.8, PyTorch 1.5, CUDA 10.2, scikit-learn 0.23, pandas 1.0.5, numpy 1.19, and were run on an updated Arch Linux System.

**Supplementary Results**

Effect of Gaussian Noise

Gaussian noise introduced into MLP_federated_ led to decreased average performance at all hospitals for predicting AKI_3_ (Supplementary Figure S2). For predicting AKI_7_, all sites had lower AUC-ROCs after the introduction of Gaussian noise (Supplemental Figure S3). These results collectively demonstrate that noise may be inserted to increase data security, but the level of noise may be fine-tuned to maintain federated model performance.

Model Performance

Additional performance metrics for all models (area under the precision recall curve, sensitivity, specificity, F1 score, accuracy) are described in Supplementary Table S4.

### **Supplementary Figures**

**Figure S1. Criteria for patient inclusion.**

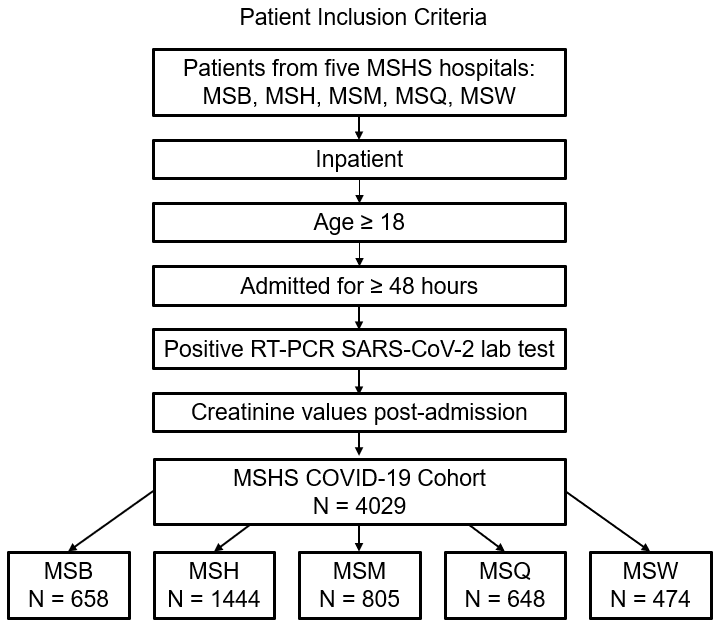

**Figure S2. Effect of Noise on Federated MLP Performance by Site for AKI_3_ Prediction.**

Performance of all models (LR_local_, LR_pooled_, LR_federated,_ LASSO_local_, LASSO_pooled_, LASSO_federated_, MLP_local_, MLP_pooled_, MLP_federated_ (no noise) by area under the receiver-operating characteristic (AUROC) at (A) Mount Sinai Brooklyn (MSB) (n=658) (B) Mount Sinai Hospital (MSH) (n=1444), (C) Mount Sinai Morningside (MSM) (n=805), (D) Mount Sinai Queens (MSQ) (n=648), and (E) Mount Sinai West (MSW) (n=474) to predict AKI within three days of admission. Averages of receiver-operating characteristic after 70-30 train-test split over 100 experiments with 95% confidence intervals are shown. Average performance of each model across all five sites is presented in (F).

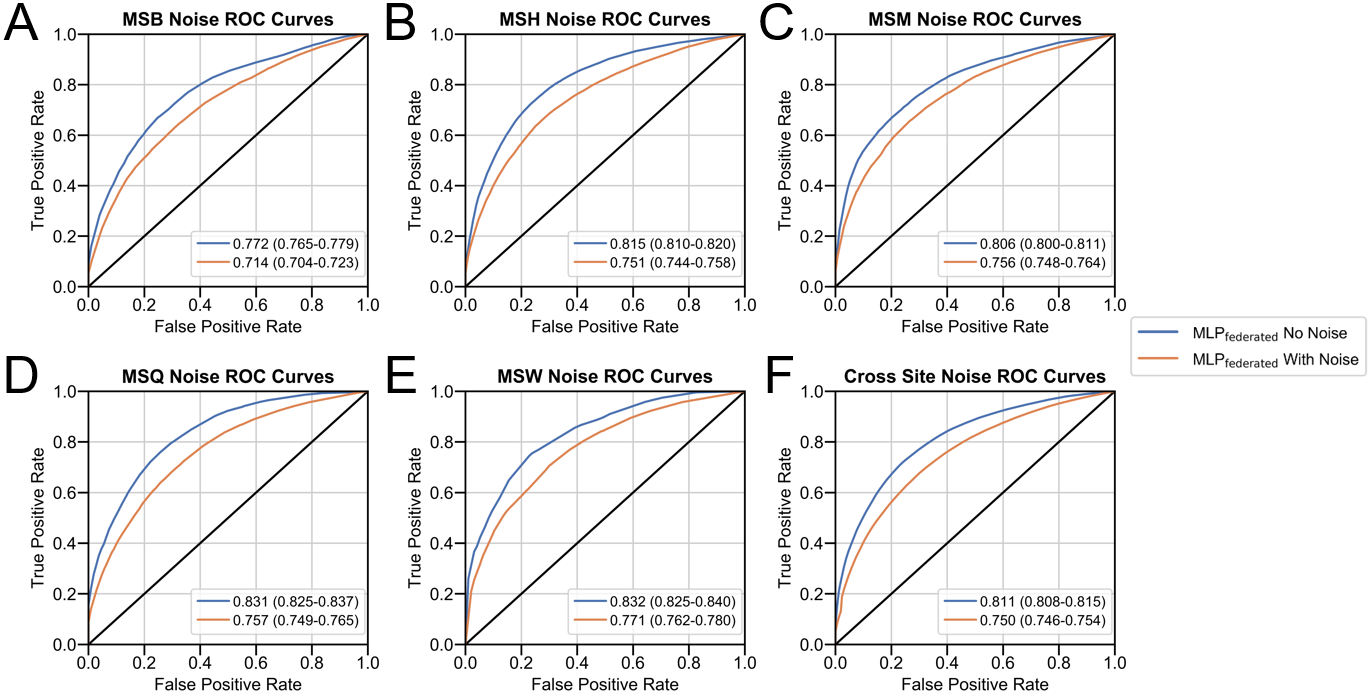

**Figure S3. Effect of Noise on Federated MLP Model Performance by Site for AKI_7_ Prediction.**

Performance of all models (LASSO_local_, LASSO_pooled_, LASSO_federated_, MLP_local_, MLP_pooled_, MLP_federated_ (no noise) by area under the receiver-operating characteristic (AUROC) at (A) Mount Sinai Brooklyn (MSB) (n=658) (B) Mount Sinai Hospital (MSH) (n=1444), (C) Mount Sinai Morningside (MSM) (n=805), (D) Mount Sinai Queens (MSQ) (n=648), and (E) Mount Sinai West (MSW) (n=474) to predict AKI within seven days of admission. Averages of receiver-operating characteristic after 70-30 train-test split over 100 experiments with 95% confidence intervals are shown. Average performance of each model across all five sites is presented in (F).

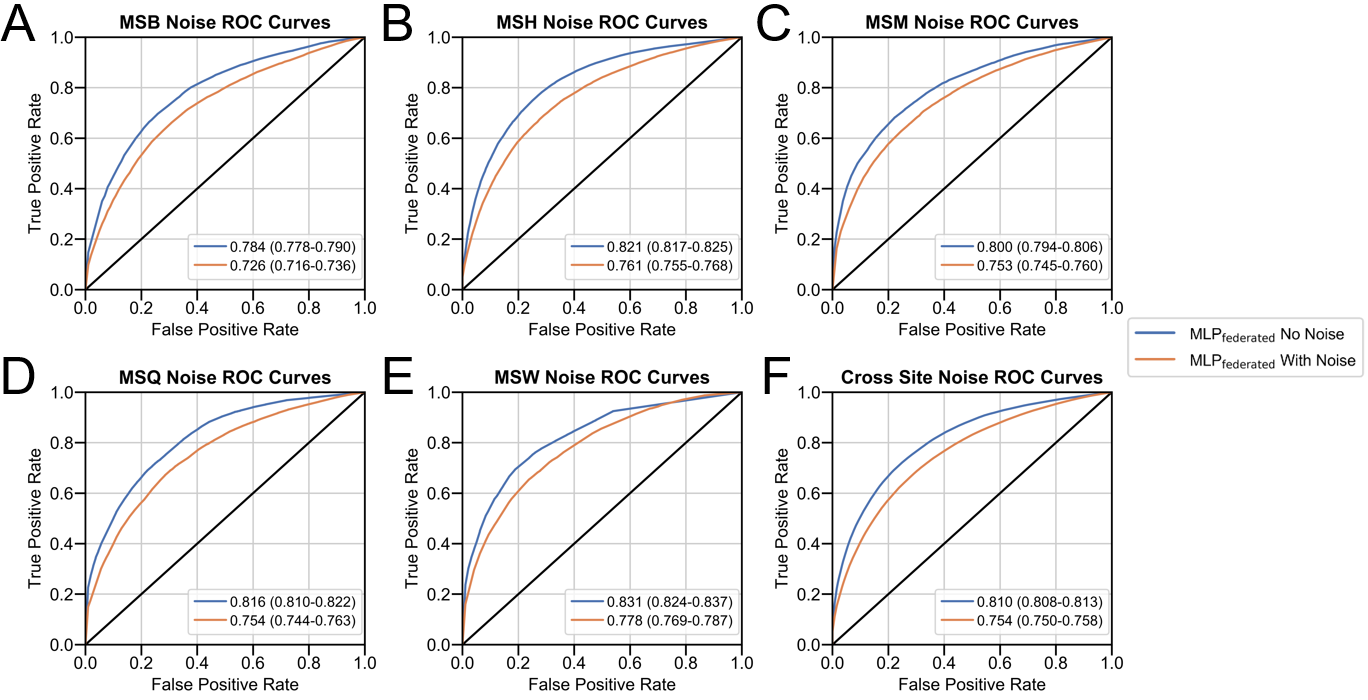

### **Supplementary Tables**

**Supplementary Table S1. TRIPOD Guidelines Report.**

| **Section/Topic** | **Item** | **Checklist Item** | **Section** |
| --- | --- | --- | --- |
| **Title and abstract** | | | |
| Title | 1 | Identify the study as developing and/or validating a multivariable prediction model, the target population, and the outcome to be predicted. | Title |
| Abstract | 2 | Provide a summary of objectives, study design, setting, participants, sample size, predictors, outcome, statistical analysis, results, and conclusions. | Abstract |
| **Introduction** | | | |
| Background and objectives | 3a | Explain the medical context (including whether diagnostic or prognostic) and rationale for developing or validating the multivariable prediction model, including references to existing models. | Introduction |
|  | 3b | Specify the objectives, including whether the study describes the development or validation of the model or both. |  |
| **Methods** | | | |
| Source of data | 4a | Describe the study design or source of data (e.g., randomized trial, cohort, or registry data), separately for the development and validation data sets, if applicable. | Short Methods |
|  | 4b | Specify the key study dates, including start of accrual; end of accrual; and, if applicable, end of follow-up. |  |
| Participants | 5a | Specify key elements of the study setting (e.g., primary care, secondary care, general population) including number and location of centers. |  |
|  | 5b | Describe eligibility criteria for participants. |  |
|  | 5c | Give details of treatments received, if relevant. | N/A |
| Outcome | 6a | Clearly define the outcome that is predicted by the prediction model, including how and when assessed. | Short Methods |
|  | 6b | Report any actions to blind assessment of the outcome to be predicted. | N/A |
| Predictors | 7a | Clearly define all predictors used in developing or validating the multivariable prediction model, including how and when they were measured. | Detailed Methods and Table S3 |
|  | 7b | Report any actions to blind assessment of predictors for the outcome and other predictors. | N/A |
| Sample size | 8 | Explain how the study size was arrived at. | Short Methods |
| Missing data | 9 | Describe how missing data were handled (e.g., complete-case analysis, single imputation, multiple imputation) with details of any imputation method. | Detailed Methods |
| Statistical analysis methods | 10a | Describe how predictors were handled in the analyses. | Detailed Methods |
|  | 10b | Specify type of model, all model-building procedures (including any predictor selection), and method for internal validation. | Detailed Methods |
|  | 10d | Specify all measures used to assess model performance and, if relevant, to compare multiple models. | Detailed Methods |
| Risk groups | 11 | Provide details on how risk groups were created, if done. | N/A |
| **Results** | | | |
| Participants | 13a | Describe the flow of participants through the study, including the number of participants with and without the outcome and, if applicable, a summary of the follow-up time. A diagram may be helpful. | Table S3 |
|  | 13b | Describe the characteristics of the participants (basic demographics, clinical features, available predictors), including the number of participants with missing data for predictors and outcome. | Table S3 |
| Model development | 14a | Specify the number of participants and outcome events in each analysis. | Table S3 |
|  | 14b | If done, report the unadjusted association between each candidate predictor and outcome. | N/A |
| Model specification | 15a | Present the full prediction model to allow predictions for individuals (i.e., all regression coefficients, and model intercept or baseline survival at a given time point). | Can’t be used directly |
|  | 15b | Explain how to the use the prediction model. | Can’t be used directly |
| Model performance | 16 | Report performance measures (with CIs) for the prediction model. | Table S4 |
| **Discussion** | | | |
| Limitations | 18 | Discuss any limitations of the study (such as nonrepresentative sample, few events per predictor, missing data). | Discussion |
| Interpretation | 19b | Give an overall interpretation of the results, considering objectives, limitations, and results from similar studies, and other relevant evidence. |  |
| Implications | 20 | Discuss the potential clinical use of the model and implications for future research. |  |
| **Other information** | | | |
| Supplementary information | 21 | Provide information about the availability of supplementary resources, such as study protocol, Web calculator, and data sets. | This file |
| Funding | 22 | Give the source of funding and the role of the funders for the present study. | Sources of Support |

**Supplementary Table S2. Final Model Hyperparameters**

| **Hyper-parameter** | **LASSO Models** | **MLP Models** |
| --- | --- | --- |
| **Penalty** | L1 |  |
| **C-value** | 0.1 |  |
| **Hidden Layers and Units** |  | Hidden layer 1: 40 units  Hidden layer 2: 10 units  Hidden layer 3: 2 units  Output layer: 1 unit |
| **Activation Function** |  | Rectified Linear Unit |
| **Optimization Function** |  | Adam |
| **Loss Function** |  | Logarithmic Softmax |
| **Dropout** |  | None |
| **Batch Size** | 32 | 32 |
| **Learning Rate** | 0.001 | 0.001 |
| **Epochs per Round** | 80 | 80 |

**Supplementary Table S3. Clinical Characteristics of Hospitalized COVID-19 Patients at Baseline.**

Clinical characteristics for all patients (n=4029) included in study with breakdown by demographics, comorbidities, vital signs, metabolic markers, liver function, inflammatory markers, hematological markers. All laboratory data was obtained within 48 hours of admission. Inter-hospital comparisons for categorical data were assessed with chi-square tests and numerical data using Kruskal-Wallis test with Bonferroni-adjusted P-values reported.

Provided as an Excel file – Supplementary_table_3.xlsx

**Supplementary Table S4. Model Performance Metrics Across Sites.**

Performance of all LASSO and MLP models (local, pooled, federated) as measured by area under the receiver operating-characteristic (AUROC), area under the precision-recall curve (AUPRC), accuracy (ACC), sensitivity (SENS), specificity (SPEC), and F1-score (F1S) with 95% confidence intervals.

Provided as an Excel file – Supplementary_table_4.xlsx

**Supplementary Table 5. Model Performance by AUROC by Hospital.**

Performance of local, pooled and federated LASSO and MLP models at each site as measured by area under the receiver operating characteristic (AUROC) curve with 95% confidence intervals.

Provided as an Excel file – Supplementary_table_5.xlsx
